# Effectiveness of CoronaVac, ChAdOx1, BNT162b2 and Ad26.COV2.S among individuals with prior SARS-CoV-2 infection in Brazil

**DOI:** 10.1101/2021.12.21.21268058

**Authors:** Thiago Cerqueira-Silva, Jason R. Andrews, Viviane S. Boaventura, Otavio T. Ranzani, Vinicius de Araújo Oliveira, Enny S Paixão, Juracy Bertoldo Júnior, Tales Mota Machado, Matt D.T. Hitchings, Murilo Dorion, Margaret L Lind, Gerson O. Penna, Derek A.T. Cummings, Natalie E Dean, Guilherme Loureiro Werneck, Neil Pearce, Mauricio L. Barreto, Albert I. Ko, Julio Croda, Manoel Barral-Netto

**Affiliations:** Instituto Gonçalo Moniz, Fiocruz, Salvador, BA, Brazil; Universidade Federal da Bahia, Salvador, BA, Brazil; Division of Infectious Diseases and Geographic Medicine, Stanford University, Stanford, CA,USA; Barcelona Institute for Global Health, ISGlobal, Barcelona, Spain; Pulmonary Division, Heart Institute, Hospital das Clínicas, Faculdade de Medicina, São Paulo, SP, Brazil; Center for Data and Knowledge Integration for Health - Fiocruz, Salvador, BA, Brazil; London School of Hygiene and Tropical Medicine, London, United Kingdom; Universidade Federal de Ouro Preto, Ouro Preto, MG, Brazil; Department of Biostatistics, College of Public Health & Health Professions, University of Florida, Gainesville, FL, USA; Department of Epidemiology of Microbial Diseases, Yale School of Public Health, New Heaven, CT, USA; Núcleo de Medicina Tropical, Universidade de Brasília, Brasília, DF, Brazil; Escola Fiocruz de Governo, Fiocruz Brasília. Brasília, DF, Brazil; Department of Biology, University of Florida, Gainesville, FL, USA; Emerging Pathogens Institute, University of Florida, Gainesville, FL, USA; Department of Biostatistics & Bioinformatics, Rollins School of Public Health, Emory University; Universidade do Estado do Rio de Janeiro, Rio de Janeiro, RJ, Brazil; Universidade Federal de Mato Grosso do Sul, Campo Grande, MS, Brazil; Fiocruz Mato Grosso do Sul, Fundação Oswaldo Cruz, Campo Grande, MS, Brazil

**Keywords:** Covid-19, CoronaVac, ChAdOx1, BNT162b2 and Ad26.COV2.S, test-negative study, case-control study, previous infections, Brazil

## Abstract

**Background:** COVID-19 vaccines have proven highly effective among SARS-CoV-2 naive individuals, but their effectiveness in preventing symptomatic infection and severe outcomes among individuals with prior infection is less clear.

**Methods:** Utilizing national COVID-19 notification, hospitalization, and vaccination datasets from Brazil, we performed a case-control study using a test-negative design to assess the effectiveness of four vaccines (CoronaVac, ChAdOx1, Ad26.COV2.S and BNT162b2) among individuals with laboratory-confirmed prior SARS-CoV-2 infection. We matched RT-PCR positive, symptomatic COVID-19 cases with RT-PCR-negative controls presenting with symptomatic illnesses, restricting both groups to tests performed at least 90 days after an initial infection. We used multivariable conditional logistic regression to compare the odds of test positivity, and the odds of hospitalization or death due to COVID-19, according to vaccination status and time since first or second dose of vaccines.

**Findings:** Among individuals with prior SARS-CoV-2 infection, vaccine effectiveness against symptomatic infection ≥ 14 days from vaccine series completion was 39.4% (95% CI 36.1-42.6) for CoronaVac, 56.0% (95% CI 51.4-60.2) for ChAdOx1, 44.0% (95% CI 31.5-54.2) for Ad26.COV2.S, and 64.8% (95% CI 54.9-72.4) for BNT162b2. For the two-dose vaccine series (CoronaVac, ChAdOx1, and BNT162b2), effectiveness against symptomatic infection was significantly greater after the second dose compared with the first dose. Effectiveness against hospitalization or death ≥ 14 days from vaccine series completion was 81.3% (95% CI 75.3-85.8) for CoronaVac, 89.9% (95% CI 83.5-93.8) for ChAdOx1, 57.7% (95% CI −2.6-82.5) for Ad26.COV2.S, and 89.7% (95% CI 54.3-97.7) for BNT162b2.

**Interpretation:** All four vaccines conferred additional protection against symptomatic infections and severe outcomes among individuals with previous SARS-CoV-2 infection. Provision of a full vaccine series to individuals following recovery from COVID-19 may reduce morbidity and mortality.

**Funding:** Brazilian National Research Council, Fundação Carlos Chagas Filho de Amparo à Pesquisa do Estado do Rio de Janeiro, Oswaldo Cruz Foundation, JBS S.A., Instituto de Salud Carlos III, Spanish Ministry of Science and Innovation, Generalitat de Catalunya.

**RESEARCH IN CONTEXT:** *Evidence before this study:* We searched PubMed, medRxiv, and SSRN for articles published from January 1, 2020 until December 15, 2021, with no language restrictions, using the search terms “vaccine effectiveness” AND “previous*” AND (“SARS-CoV-2” OR “COVID-19”). We found several studies evaluating ChAdOx1 and BNT162b2, and one additionally reporting on mRNA-1273 and Ad26.COV2.S, which found that previously infected individuals who were vaccinated had lower risk of symptomatic SARS-CoV-2 infection. One study found that risk of hospitalization was lower for previously infected individuals after a full series of BNT162b2 or mRNA-1273. Limited evidence is available comparing effectiveness of one versus two doses among individuals with prior infection. No studies reported effectiveness of inactivated vaccines among previously infected individuals.

*Added value of this study:* We used national databases of COVID-19 case surveillance, laboratory testing, and vaccination from Brazil to investigate effectiveness of CoronaVac, ChAdOx1, Ad26.COV2.S and BNT162b2 among individuals with a prior, laboratory-confirmed SARS-CoV-2 infection. We matched >22,000 RT-PCR-confirmed re-infections with >145,000 RT-PCR-negative controls using a test-negative design. All four vaccines were effective against symptomatic SARS-CoV-2 infections, with effectiveness from 14 days after series completion ranging from 39-65%. For vaccines with two-dose regimens, the second dose provided significantly increased effectiveness compared with one dose. Effectiveness against COVID-19-associated hospitalization or death from 14 days after series completion was >80% for CoronaVac, ChAdOx1and BNT162b2.

*Implications of all the available evidence:* We find evidence that four vaccines, using three different platforms, all provide protection to previously infected individuals against symptomatic SARS-CoV-2 infection and severe outcomes, with a second dose conferring significant additional benefits. These results support the provision of a full vaccine series among individuals with prior SARS-CoV-2 infection.

## INTRODUCTION

Over 250 million confirmed cases of COVID-19 have been reported since the start of the pandemic,^1^ and the true cumulative incidence has likely been several times greater. Within a year of the identification of SARS-CoV-2, multiple vaccines were developed, found to be highly efficacious among seronegative individuals in clinical trials and introduced into national vaccination programs.^2–5^ Uptake of COVID-19 vaccines has been variable across populations amid hesitancy, and public debate has emerged about the need for vaccination among people who have had previous SARS-CoV-2 infection,^6^ and if so, whether a single dose is sufficient.^7,8^ The emergence of more transmissible variants with enhanced immune escape, and resulting waves of infection and reinfection, have renewed questions about the importance of vaccination in individuals with prior COVID-19.^9,10^

SARS-CoV-2 infection induces robust T-cell and B-cell responses,^11^ and the risk of symptomatic infection and severe outcomes is lower among people with prior COVID-19 infection compared with naive individuals.^12^ Emerging evidence suggests that vaccination with ChAdOx1, Ad26.COV2.S, BNT162b2, or mRNA-1273 does confer additional protection against symptomatic reinfection among individuals with prior SARS-CoV-2 infection.^13–19^ However, only one study has assessed protection against severe outcomes, with just 75 hospitalizations and 2 deaths.^19^ Moreover, data for inactivated vaccines, which account for almost half of all doses given globally, are lacking.^20^

Brazil has recorded more than 22 million COVID-19 infections and 600,000 deaths to date. In January 2021, a national COVID-19 immunization program was initiated, which has utilized four vaccines of three different classes: inactivated (CoronaVac; Sinovac), viral vector (ChAdOx1; AstraZeneca and Ad26.COV2.S; Janssen) and mRNA (BNT162b2; Pfizer-BioNTech). We utilized national disease surveillance and vaccination databases to estimate the effectiveness of these four vaccines, among individuals with laboratory-confirmed previous infections, against symptomatic infection, hospitalization, and death.

## METHODS

### Study design, population, and data sources

We conducted a test negative design (TND) case-control study to evaluate effectiveness of four vaccines (CoronaVac, ChAdOx1, Ad26.COV2.S and BNT162b2) in individuals with prior SARS-CoV-2 infection in Brazil. The study population included individuals with a prior positive reverse transcriptase polymerase chain reaction (RT-PCR) or rapid antigen test for SARS-CoV-2 who presented again to health facilities with symptomatic illness and were tested for SARS-CoV-2 at least 90 days after their first positive test. We matched individuals who tested positive on these subsequent test (cases) to those who tested negative (controls).

We utilized data from several national data sources: a deterministically linked dataset comprised of the Programa Nacional de Imunizações (PNI), which contains records of all vaccines administered in Brazil; the e-SUS Notifica, which contains records of suspected and confirmed COVID-19 in outpatient clinics; and the Sistema de Informação da Vigilância Epidemiológica da Gripe (SIVEP-Gripe), which contains records of COVID-19 hospitalizations and deaths. All data were pseudo-anonymized with a common unique identifier provided by the Brazilian Ministry of Health. The research protocol was approved by the Brazilian National Commission in Research Ethics (CONEP) (approval number 4.921.308).

Brazil’s national COVID-19 immunization program commenced on January 17, 2021. Rollout plans were determined at state and local levels; healthcare workers and elderly individuals were the first groups eligible, with age criteria for eligibility advancing downwards with calendar time. Four vaccines have been offered in immunization programs in Brazil: 1) CoronaVac (Sinovac), provided as a two-dose series with a 4-week interval between doses; 2) ChAdOx1 (AstraZeneca), provided as a two-dose series with 12-week between doses which was subsequently reduced to 8 weeks in some states; 3) Ad26.COV2.S (Janssen) as a single dose series; and 4) BNT162b2 (Pfizer-BioNTech), as a two-dose series, initially with a 12-week interval, which was subsequently reduced to 3 weeks in some states.

### Eligibility and selection of cases and controls

Inclusion criteria for this study included: 1) age ≥18 years; 2) prior SARS-CoV-2 infection confirmed by RT-PCR or rapid antigen test; 3) a second exam (RT-PCR test) fulfilling the following criteria: a) associated with an event of symptomatic illness and occurring within 10 days of symptom onset; b) at least 90 days after their first positive test; and c) occurring after the vaccination program began in Brazil (January 17, 2021). We included individuals with first infection between February 24, 2020, and August 13, 2021, and second RT-PCR test occurring between January 18, 2021, and November 11, 2021.

We excluded: 1) individuals for whom data were incomplete on age, sex, location of residence, vaccination status, testing status or dates; 2) individuals who received a different vaccine for the second dose from the first; 3) individuals whose time interval between the first and second doses was less than 14 days; 4) individuals vaccinated before the first infection or less than 14 days after the first infection. For tests, we excluded: 1) negative tests followed by a positive test within 7 days (to avoid misclassification); 2) tests occurring after the second positive test; 3) tests with symptom onset date occurring after the notification of suspected case in the surveillance system; 4) tests occurring among individuals lacking symptoms; 5) tests occurring after a 3rd dose of vaccine, as we were not powered to examine effectiveness of third doses in this analysis.

We matched cases, defined as eligible SARS-CoV-2 RT-PCR tests that were positive, with controls, defined as eligible SARS-CoV-2 RT-PCR tests that were negative. Hospitalization or death related to COVID-19 was defined by a positive SARS-CoV-2 RT-PCR test accompanied by hospital admission or death occurring within 28 days of the sample collection date. For each outcome, we selected matched pairs in which cases had the outcome of interest, and fit the model described above to each subset. For severe outcomes, controls therefore represented test-negative patients from ambulatory or hospital settings who had RT-PCR testing, to reflect the population at risk for that outcome. We matched one case to a maximum of ten controls, with replacement, by date of RT-PCR testing (±10 days), age (±5 years), sex, and municipality of residence. Individuals who were selected as cases could also serve as controls if they had negative tests that were collected >7 days before their positive test.

### Statistical analyses

The primary outcomes of interest were symptomatic SARS-CoV-2 re-infection and hospitalization or death following SARS-CoV-2 infection. The primary exposure of interest was vaccination status, which was categorized by vaccine and according to the status at the time of RT-PCR test collection as unvaccinated; 0-13 days post first dose; ≥14 days post first dose; 0-13 days post second dose; or ≥14 days post second dose. Post-dose 2 is not applicable to Ad26.COV2.S. We considered vaccine effectiveness against symptomatic SARS-CoV-2 infection and against COVID-19-related hospitalization or death, among individuals with prior confirmed SARS-CoV-2 infection, ≥14 days after series completion (after 2 doses of CoronaVac, ChAdOx1, and BNT162b2 and after one dose of Ad26.COV2.S) to be the primary estimands of interest. We considered effectiveness in the 6 days after the first dose as an indicator of bias, as we expected, protection to be minimal during this time and substantial differences in risk could reflect residual confounding between the vaccinated and unvaccinated populations.^21^

We estimated vaccine effectiveness (1-Odds Ratio) using conditional logistic regression, accounting for the matched design, with vaccination status (including number of doses and time period since dose) as the predictor and adjusting for the number of reported chronic comorbidities (categorized as none, one, and at least two), pregnancy, postpartum period, self-reported race, days elapsed between first positive test and second test (as a restricted cubic spline), and whether the individual was hospitalized during their first SARS-CoV-2 infection, for severe outcomes age (as continuous) was also included due to residual confounding.

We performed subgroup analyses in which we assessed vaccine effectiveness restricted to individuals above and below 50 years of age, among individuals who had completed their vaccine series greater or less than 90 days prior (to assess for possible waning), and among individuals tested at least 180 days after their initial positive test. Generalized linear hypothesis tests were used for comparisons across different vaccination status, and the confidence intervals and *p*-values were not adjusted for multiple comparisons. All data processing and analyses were performed in R (version 4.1.1), using the following packages: *tidyverse, multcomp, MatchIt* and *survival*.^22–25^

## RESULTS

Brazil had two COVID-19 epidemic waves, with the first occurring between July and September 2020, and the second between February and June 2021, during which the Gamma variant was dominant (**Figure 1**). Brazil’s national vaccination program commenced on January 17, 2021; fifty percent of the adult population had received a first vaccine dose by July 07, 2021. Between February 24, 2020, and November 11, 2021, there were more than 23 million individuals with valid SARS-CoV-2 tests and 11 million confirmed cases (**Figure 2**). Among these, we identified 241,845 individuals who had a subsequent, symptomatic illness with RT-PCR testing performed at least 90 days after their initial SARS-CoV-2 infection and after the vaccination program commenced. Among these, 30,910 (14.5%) had positive RT-PCR. We matched 22,566 of these cases with 145,055 negative RT-PCR tests from 68,426 individuals as controls. Among cases, 1,545 (6.8%) were hospitalized and 290 (1.3%) died within 28 days of a positive SARS-CoV-2 RT-PCR.

**Figure 1.**
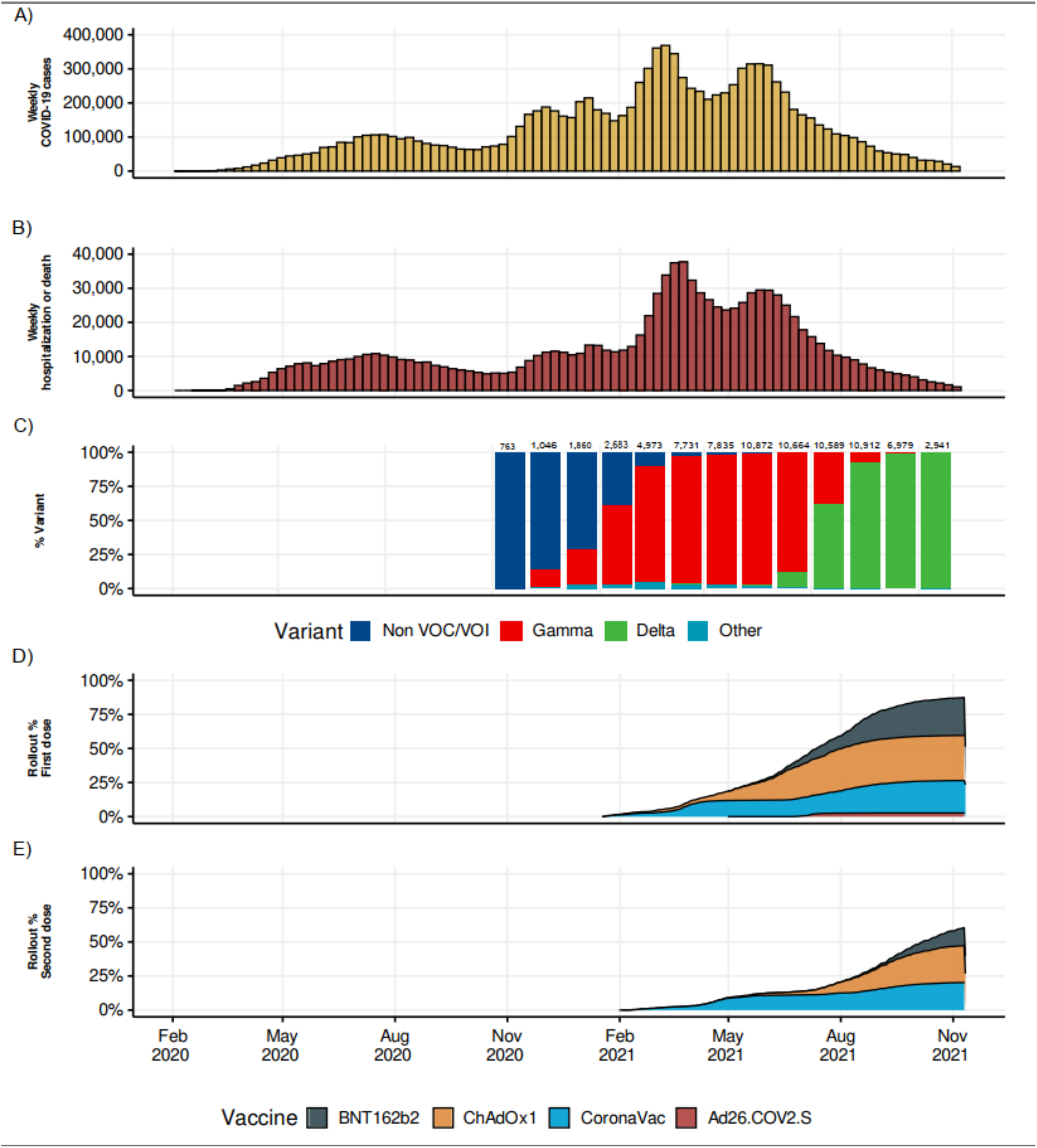
Temporal trends in COVID-19 cases, hospitalization or deaths, variants and vaccination coverage from national databases in Brazil. (A) Weekly numbers of symptomatic COVID-19 cases, (B) COVID-19 associated hospitalizations or deaths reported in national databases; (C) monthly proportions of variants of concern among sequenced SARS-CoV-2 (number of sequenced viruses are shown above each bar); and cumulative proportion of the population over 11 years of age who received a first (D) or second(E) dose of each vaccine.

**Figure 2.**
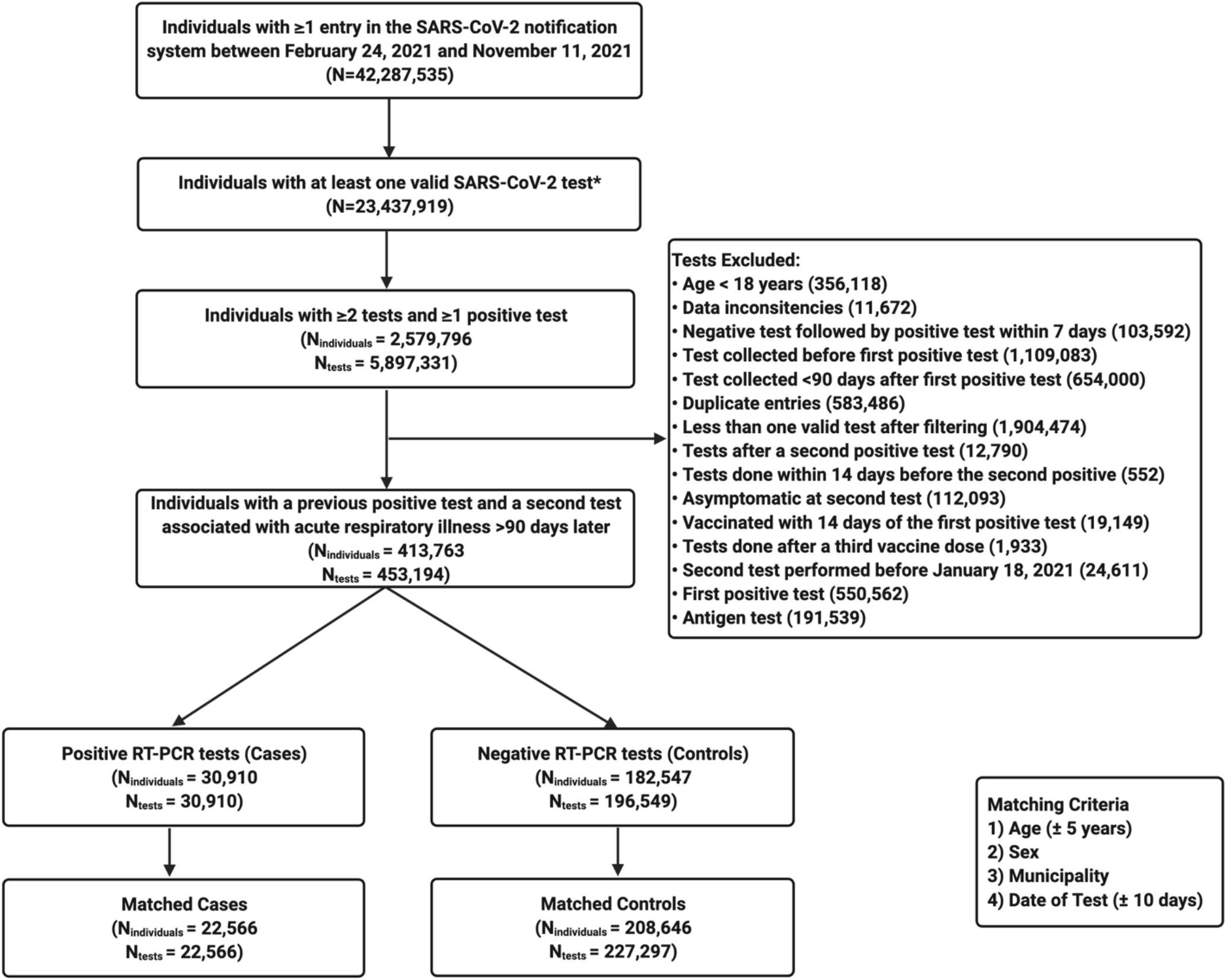
Flowchart of the study population from surveillance databases and selection of matched cases and controls. *Antigen or RT-PCR collected less or equal than 10 days of symptom onset. ^1^Some individuals contributed as controls and cases, and matching allowed for replacement of controls between cases. RT-PCR: reverse transcription polymerase chain reaction

Demographics and clinical characteristics of eligible and matched pairs are presented in Table 1. The median age of the matched population was 36 (interquartile range [IQR]: 29-44) years, approximately 60% of cases and controls were female, and the median time between first infection and the subsequent RT-PCR test was similar between the cases (216 days, IQR 146-291) and controls (223 days, IQR 154-295). Most cases (64.5%) and controls (57.4%) were unvaccinated at the time of the test. Among individuals vaccinated, 17,008 (42.8%) received CoronaVac, 15,897 (40.1%) received ChAdOx1, 5,935 (14.9%) received BNT162b2 and 877 (2.2%) received Ad26.COV2.S. Demographic characteristics were similar among vaccine recipients included in the analysis, but recipients of ChAdOx1 tended to be older and have more comorbidities (**Appendix, Table A1**). The median time between vaccination and test was 34 days (IQR 17-61) for individuals with only one dose and 59 (IQR 27-105) for individuals with two doses, which differed by each vaccine (**Appendix, Figure A3**).

**Table 1.** Characteristics of individuals eligible for and matched into case-test negative pairs.

| Characteristic | Eligible cases and controls |  | Matched pairs |  |
| --- | --- | --- | --- | --- |
|  | Case<br>N = 30,910 | Control<br>N = 196,549 | Case<br>N = 22,566 | Control<br>N = 145,055 |
| Individuals | 30,910 (100) | 182,547 (92.9) | 22,566 (100) | 68,426 (47.2) |
| Age in years, median (IQR) | 38 (29 – 47) | 37 (28 – 47) | 37 (29 – 46) | 36 (29 – 44) |
| Sex, female | 18,106 (58.6) | 119,134 (60.6) | 13,631 (60.4) | 90,931 (62.7) |
| Race |  |  |  |  |
| White | 13,841 (44.8) | 109,923 (55.9) | 10,302 (45.7) | 67,403 (46.5) |
| Mixed | 11,363 (36.8) | 53,401 (27.2) | 7,998 (35.4) | 50,788 (35.0) |
| Black | 1,420 (4.6) | 9,034 (4.6) | 1,052 (4.7) | 7,572 (5.2) |
| Indigenous or Asian | 2,081 (6.7) | 9,305 (4.7) | 1,437 (6.4) | 8,751 (6.0) |
| Missing | 2,205 (7.1) | 14,886 (7.6) | 1,777 (7.9) | 10,541 (7.3) |
| Region of residence |  |  |  |  |
| Central-west | 3,260 (10.5) | 46,968 (23.9) | 2,302 (10.2) | 12,997 (9.0) |
| North | 2,406 (7.8) | 9,724 (4.9) | 1,870 (8.3) | 12,372 (8.5) |
| Northeast | 8,268 (26.7) | 30,027 (15.3) | 5,297 (23.5) | 30,489 (21.0) |
| South | 2,823 (9.1) | 16,251 (8.3) | 1,991 (8.8) | 14,745 (10.2) |
| Southeast | 14,153 (45.8) | 93,579 (47.6) | 11,106 (49.2) | 74,452 (51.3) |
| Residence in state capital | 9,250 (29.9) | 51,128 (26.0) | 8,982 (39.8) | 77,198 (53.2) |
| Medical comorbidities |  |  |  |  |
| 0 | 25,988 (84.1) | 166,655 (84.8) | 19,271 (85.4) | 124,964 (86.1) |
| 1 | 3,552 (11.5) | 22,178 (11.3) | 2,459 (10.9) | 15,360 (10.6) |
| ≥2 | 1,370 (4.4) | 7,716 (3.9) | 836 (3.7) | 4,731 (3.3) |
| Days from first positive test to second test, median (IQR) | 210 (144–285) | 217 (154–293) | 216 (146–291) | 223 (154–295) |
| Hospitalized during 1st infection | 1,220 (3.9) | 9,481 (4.8) | 781 (3.5) | 6,507 (4.5) |
| Days, 1st vaccine dose to 2nd positive test, median (IQR) | 161 (91–237) | 156 (91–231) | 168 (94–244) | 178 (101–249) |
| Hospitalization (up to 28 days) | 2,508 (8.1) | 3,770 (1.9) | 1,545 (6.8) | 2,196 (1.5) |
| Death (up to 28 days) | 559 (1.8) | 663 (0.3) | 290 (1.3) | 386 (0.3) |
| Hospitalization or death | 2,554 (8.3) | 3,829 (1.9) | 1,564 (6.9) | 2,238 (1.5) |

### Vaccine effectiveness against symptomatic SARS-CoV-2 re-infection

Effectiveness against symptomatic SARS-CoV-2 reinfection ≥ 14 days from vaccine series completion was 39.4% (95% CI 36.1-42.6) for CoronaVac, 56.0% (95% CI 51.4-60.2) for ChAdOx1, 44.0% (95% CI 31.5-54.2) for Ad26.COV2.S, and 64.8% (95% CI 54.9-72.4) for BNT162b2 (**Figure 3**). The two-dose vaccines (CoronaVac, ChAdOx1, and BNT162b2) all showed a significant increase in protection from ≥14 days after the first dose, to 0-13 days and ≥14 days after the second dose. For CoronaVac, effectiveness was twice as high in the period ≥14 days after the second dose compared with ≥14 days after the first (39.4% vs 18.8%, p<0.001). Only CoronaVac demonstrated protection (21.0%, 95% CI 2.3-36.1) against symptomatic infection within six days of the first dose, which we used as a test of bias (**Appendix, Table A2**).

**Figure 3.**
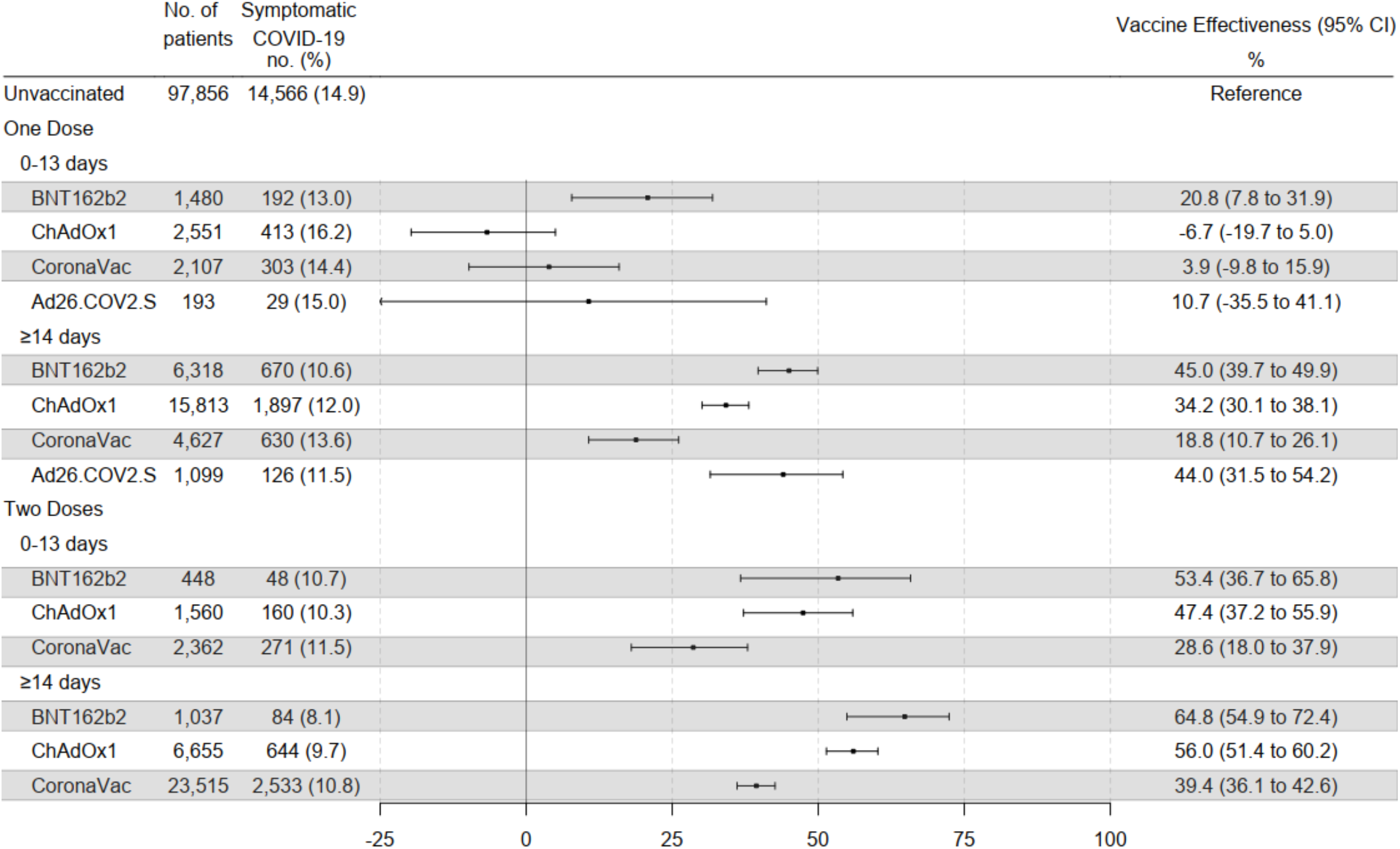
Effectiveness of the BNT162b2, ChAdOx1, CoronaVac and Ad26.COV2.S vaccines against symptomatic COVID-19 among individuals with prior SARS-CoV-2 infection.

### Vaccine effectiveness against Covid-19 related hospitalization or death

From 14 days after the completion of the series, effectiveness against COVID-19 related hospitalization or death was 81.3% (95% CI 75.3-85.8) for CoronaVac, 89.9% (95% CI 83.5-93.8) for ChAdOx1, 57.7% (95% CI −2.6-82.5) for Ad26.COV2.S, and 89.7% (95% CI 54.3-97.7) for BNT162b2 (**Figure 4**). Effectiveness ≥14 days after a single dose for was 56.9% (95% CI 45.2-66.1) for ChAdOx1 and 61.8% (95% CI 40.8-75.3) for BNT162B2, but was modest for CoronaVac (35.3%; 95% CI 7.9-54.5). Effectiveness against hospitalization or death was significantly greater ≥14 days after two doses compared to ≥14 days after one dose for CoronaVac (81.3% vs 35.3%, p<0.001) and ChAdOx1 (89.9% vs 56.9%, p<0.001) and non-significantly increased for BNT162b2 (89.7% vs 61.8%, p=0.091). We found no evidence of protection for all four vaccines against COVID-19 related hospitalization or death within 6 days of the first dose (**Appendix, Table A2**).

**Figure 4.**
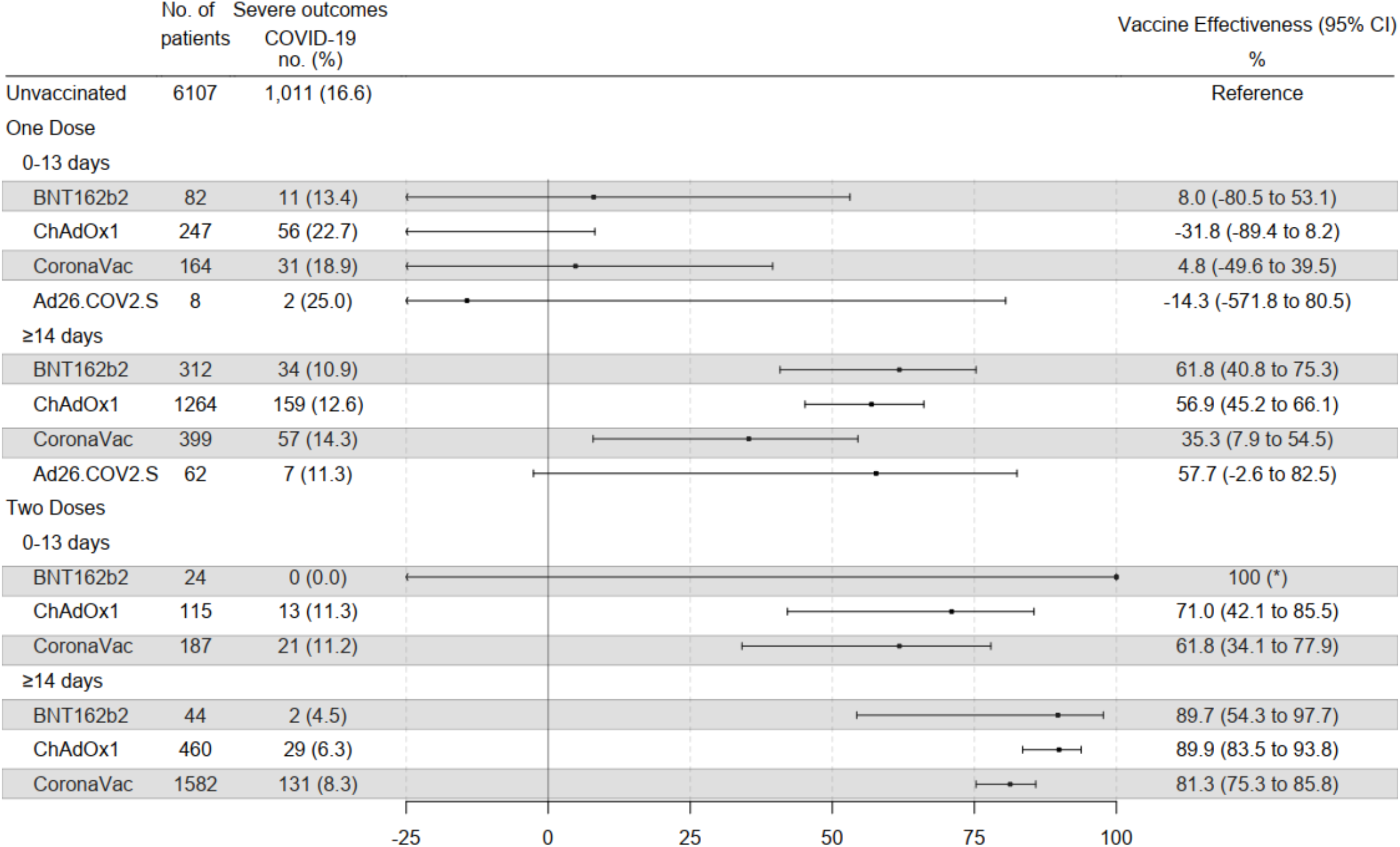
Effectiveness of the BNT162b2, ChAdOx1, CoronaVac and Ad26.COV2.S vaccines against COVID-19-associated hospitalization or death among individuals with prior SARS-CoV-2 infection.

### Vaccine effectiveness in subpopulations

For the primary estimands of vaccine effectiveness against symptomatic SARS-CoV-2 infection and against COVID-19-related hospitalization or death ≥14 days after series completion, we found no differences between age groups (≥50 years, <50 years). For three of the vaccines, we saw a trend towards increased effectiveness against symptomatic infection for vaccination given 91-180 days after prior infection compared with >180 days, including a significant increase for BNT162b2 (35.3% vs 70.7%, p=0.011) (**Appendix, Table A3**). There were no differences in effectiveness against symptomatic infection comparing the periods of ≥14-90 days and >90 days post vaccine series completion. For hospitalization and death, effectiveness of ChAdOx1 was greater >90 days post-completion compared with 14-90 (95.1% vs 86.6%; p=0.007) whereas it was lower for CoronaVac over the later interval (74.4% vs 86.6%, p=0.012) (**Appendix, Tables A3 and A4**).

## DISCUSSION

In this nationwide, population-based study among individuals with confirmed prior SARS-CoV-2 infection, we observed a high degree of additional protection of four vaccines against symptomatic COVID-19 and severe outcomes. For the three vaccines with two doses (CoronaVac, ChAdOx1, and BNT162b2), additional protection against symptomatic infection was observed after the second dose, and protection against hospitalization or death exceeded 80%. These results support vaccination, including the full vaccine series, among individuals with prior SARS-CoV-2 infection.

There has been public debate about whether previously infected individuals need to be vaccinated, due to substantial immunity conferred by SARS-CoV-2 infection.^6^ Furthermore, in view data demonstrating robust immune responses following a first vaccine dose in previously infected individuals, some have argued that two doses are not necessary.^7,8^ Indeed, several countries recommend that a single vaccine dose is sufficient for previously infected individuals.^26–28^ We found that a second dose of CoronaVac, ChAdOx1, and BNT162b2 provided significant additional protection against symptomatic infections and severe disease.

The results of this analysis are consistent with recent studies reporting that individuals with prior infection who received ChAdOx1 and BNT162b2 had a lower risk of symptomatic COVID-19 than unvaccinated, previously infected individuals.^13,14,16,17^ Direct comparison with vaccine effectiveness estimates from these is challenged by differences in design, with most studies reporting risk in comparison against unvaccinated, SARS-CoV-2-naive individuals. However, inferred protection from those studies ranged from 40-94%, consistent with the magnitude of protection against symptomatic infection found for ChAdOx1 (56.0%) and BNT162b2 (64.8%) in this study. Our analysis additionally adds new estimates on effectiveness of CoronaVac and Ad26.COV2.S vaccines among previously infected individuals, finding that they provide more modest levels of protection (39.4% and 44.0%, respectively) against symptomatic infection, consistent with their lower effectiveness in naive populations.^29,30^ Concerns have been raised about less robust and durable neutralizing antibody responses in SARS-CoV-2-naive individuals receiving CoronaVac compared with other vaccines.^31^ We found that two doses of CoronaVac provided high levels of protection against severe outcomes (81.3%, 95% CI 75.3-85.8). As CoronaVac is among the most widely used vaccines in the world, these findings have broad implications for many national programs.^20^

To our knowledge, only one prior study reported vaccine effectiveness among previously infected individuals against COVID-19 related hospitalization or death; with just 75 outcomes and three vaccines evaluated, power in that study was limited for assessing vaccine and dose-specific effectiveness, but estimates ranged from 58% (BNT162b2) to 68% (mRNA-1273), with no significant protection from Ad26.COV2.S.^19^ We found that protection against these severe outcomes, from 14 days after the second dose, was greater than 80% for the three two-dose vaccines (CoronaVac, ChAdOx1, and BNT162b2). These results are consistent with recent data showing that previously infected individuals have even greater increases in T-cell and B-cell responses following vaccination compared with those without prior infection.^32^ This high degree of hybrid immunity, from infections and vaccination,^33^ could potentially explain why Brazil, despite having similar vaccination coverage as the United States and many European countries, did not have a similar increase in hospitalizations and deaths in the period where the Delta variant become dominant.

Effectiveness against severe outcomes was lower (57.7%) for the single-dose Ad26.COV2.S vaccine compared with the vaccines given in two dose series, though the confidence limits were wide. The Ad26.COV2.S vaccine was used in a more focal rollout from June to July, 2021, and far fewer individuals receive this vaccine compared with the others. Brazil’s Ministry of Health now recommends that individuals who received this vaccine receive a second dose after 60 days.

We focused our analyses on previously infected individuals to address the question of whether and to what extent vaccines confer additional protection against symptomatic infection and severe outcomes. We did not compare against previously uninfected individuals, as their risk of exposure may be quite different, which could lead to biased estimates in this population-based study. Additionally, there is substantial risk of misclassification of previously infected individuals as not previously infected, due to incomplete surveillance and asymptomatic infections; restricting vaccine effectiveness analysis to individuals with PCR-confirmed prior infection avoids this bias. While there has been much discussion concerning the relative protection conferred by infected-derived immunity and vaccine-derived immunity, from a medical and public health standpoint, the critical question is understanding whether individuals with prior infection would benefit from vaccination. This study suggests that individuals infected before vaccination benefit from strong protection against severe outcomes with all four vaccines studied.

A major difficulty with observational studies of vaccine effectiveness is the risk of confounding, whereby differences in the vaccinated and unvaccinated populations are associated with risk of diagnosis of COVID-19. The matched, test-negative design has been recommended by the World Health Organization^34^ to mitigate risk of confounding introduced by care-seeking and diagnostic access; nevertheless, residual confounding may occur. We used vaccine effectiveness in the six days following the first dose as a bias indicator, in that differences during this period before vaccine-conferred protection is expected could indicate counfounding.^21^ We only observed significant effectiveness in this time interval for one vaccine (CoronaVac) and one outcome (symptomatic infection); interestingly, over the 7-13 day time window, no effectiveness (−9.6%, 95% CI −29.4-7.2) was observed. Whether the effectiveness observed over days 0-6 reflects bias or chance among the 8 bias indicator tests is unclear, but the absence of effects in the 7-13 day window may point against systematic differences in the CoronaVac recipients with respect to SARS-CoV-2 risk. For BNT162b2, we found substantial protection (27%, 95% CI 10 to 41%) in the 7-13 day window. In clinical trials of BNT162b2, efficacy was apparent from approximately 11 days after the first dose.^2^ Given the rapid and robust immune responses following first vaccination among previously infected individuals, we believe these findings are consistent with early vaccine-conferred immunity.

This study was subject to several limitations. First, we were not powered to assess vaccine effectiveness by age groups. We compared effectiveness in individuals above and below the age of 50 and did not observe major differences. Second, there were differences in the timing of introduction and eligibility for each of the vaccines. This should prompt some caution in the comparison of effectiveness between vaccines, as the calendar period and median duration from second dose differed somewhat between vaccines. For example, if effectiveness wanes over time, vaccines used earlier would have lower effectiveness than those introduced later. Additionally, changes in variant distribution during the study period could alter effectiveness by time since vaccination. We did not have individual level data on variants, which precluded assessment of variant-specific vaccine effectiveness. We used a matched, test-negative design with multivariable regression to reduce non-vaccine related differences between the cases and controls; however, there could be unmeasured differences that lead to confounding. Finally, an important question is when vaccines should be given to individuals with previous SARS-CoV-2 infection, and our study was unable to address this. To avoid misclassification of reinfections, we only considered tests performed at least 90 days after the initial infection. We examined whether individuals vaccinated from 91-180 days after initial infection had differential protection from those vaccinated after 180 days, and did not detect any differences. However, we could not assess longer time periods.

The accelerated development of effective vaccines against COVID-19 has been a remarkable scientific achievement, but more than 40% of the world’s population has yet to receive a first dose, and a substantial proportion of these individuals have already been infected with SARS-CoV-2. The results of this study provide evidence for the benefits of vaccination among individuals who have already been infected with SARS-CoV-2, with all four vaccines conferring substantial reductions in hospitalization and death due to COVID-19. Ensuring vaccine access to individuals with prior infection may be particularly important amid early reports of the Omicron variant, which suggest that immunity conferred by prior infection is reduced.^9,10^ Expanded, equitable rollout of vaccines for all individuals remains critical for mitigating the continued threat posed by SARS-CoV-2.

## Data Availability

All data produced in the present study are available upon reasonable request to the authors

## Acknowledgements

This study was partially supported by a donation from the “Fazer o bem faz bem” program from JBS S.A.. GLW, MLB, and JC and MB-N are research fellows from CNPq, the Brazilian National Research Council. GLW acknowledges Fundação Carlos Chagas Filho de Amparo à Pesquisa do Estado do Rio de Janeiro (E-26/210.180/2020). JC is supported by the Oswaldo Cruz Foundation (Edital Covid-19 – resposta rápida: 48111668950485). OTR is funded by a Sara Borrell fellowship (CD19/00110) from the Instituto de Salud Carlos III. OTR acknowledges support from the Spanish Ministry of Science and Innovation through the Centro de Excelencia Severo Ochoa 2019-2023 Program and from the Generalitat de Catalunya through the CERCA Program.

## Supplementary appendix

### Supplement

**Figure A1.**
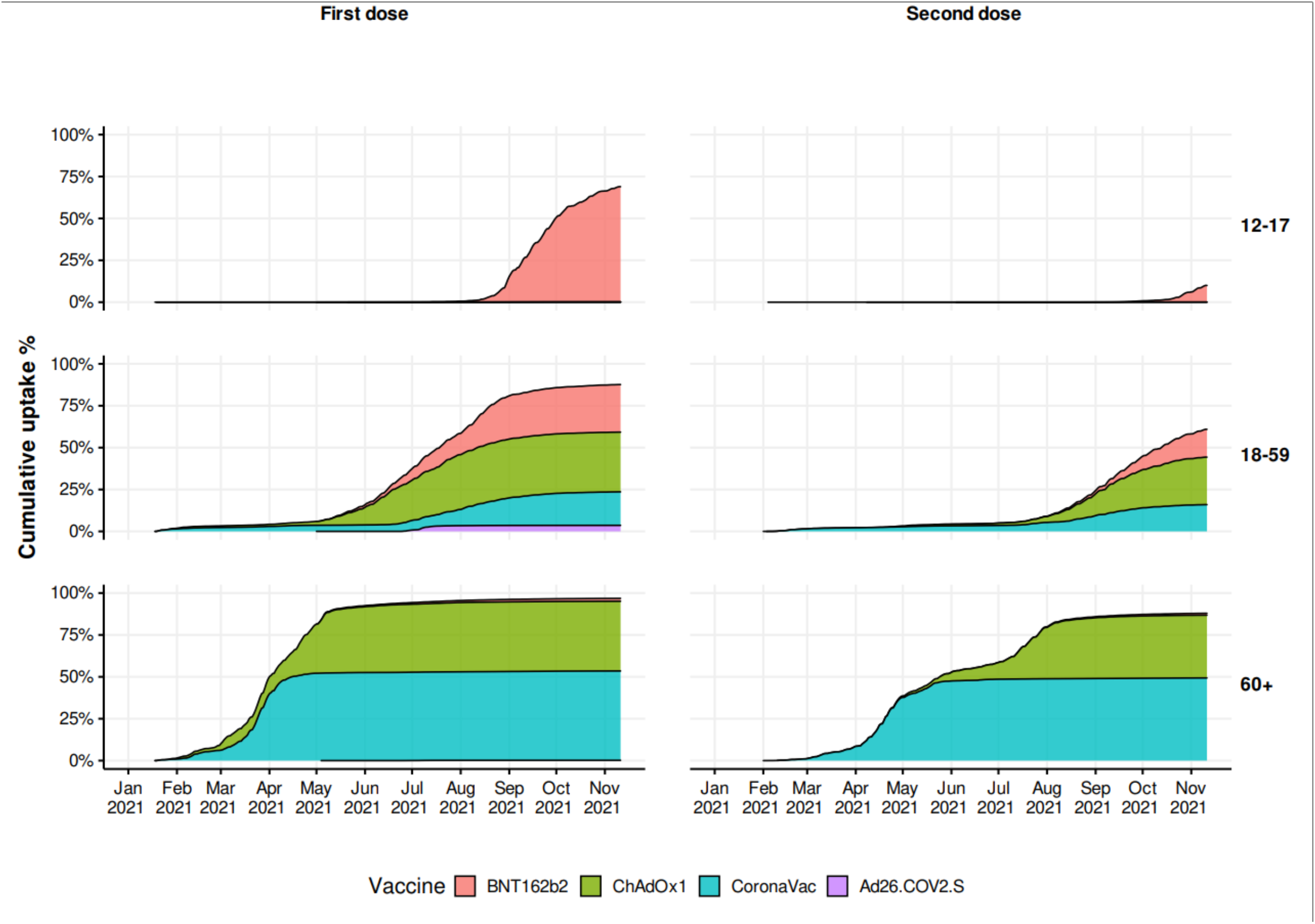
Cumulative proportion of the population receiving each vaccine, by age group.

**Figure A2.**
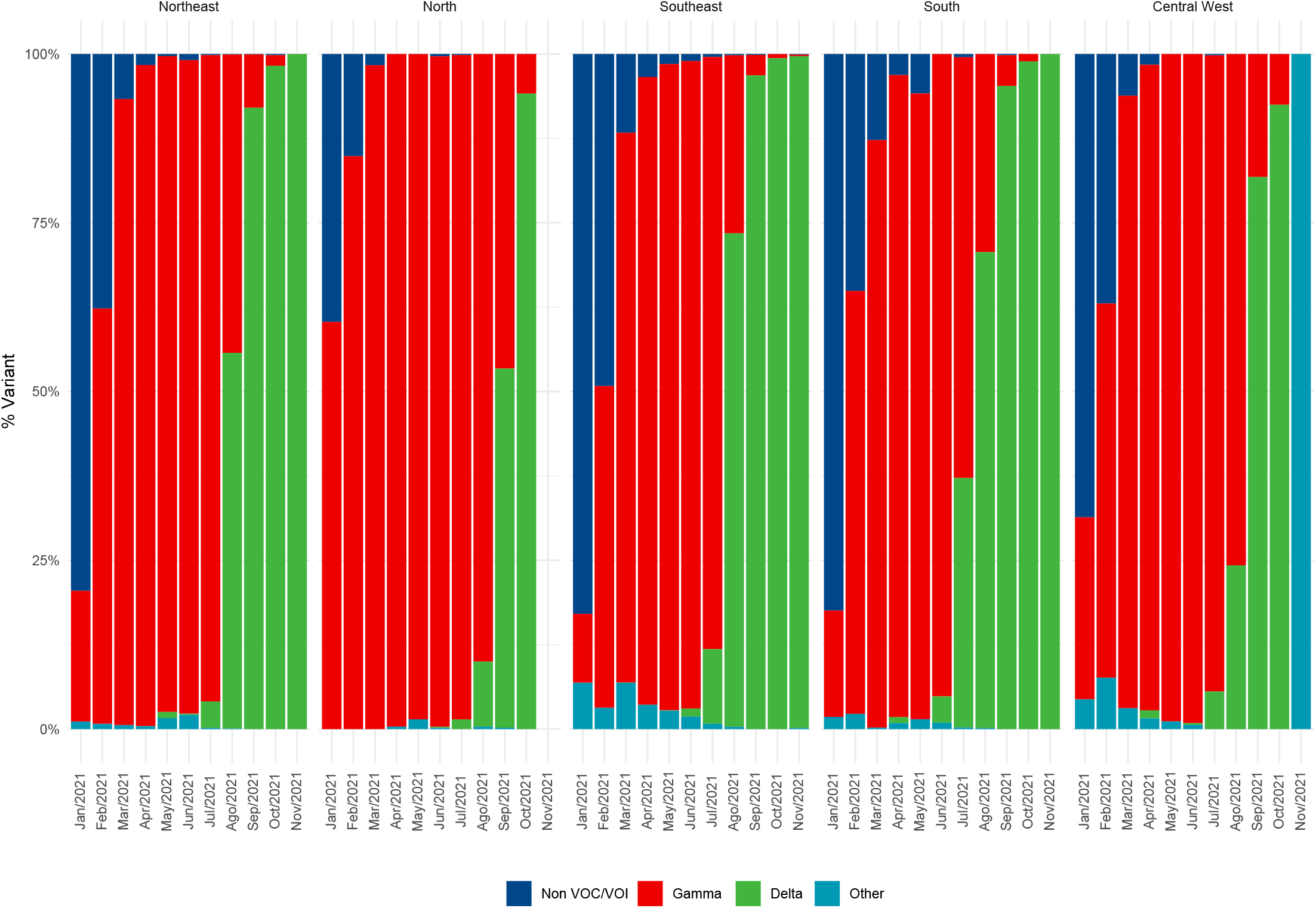
Prevalence of SARS-CoV-2 variants of concern in Brazil from January 01, 2021, to November, 30, 2021, stratified by region. Data obtained from the Fiocruz Genomic Network and GISAID.

**Figure A3.**
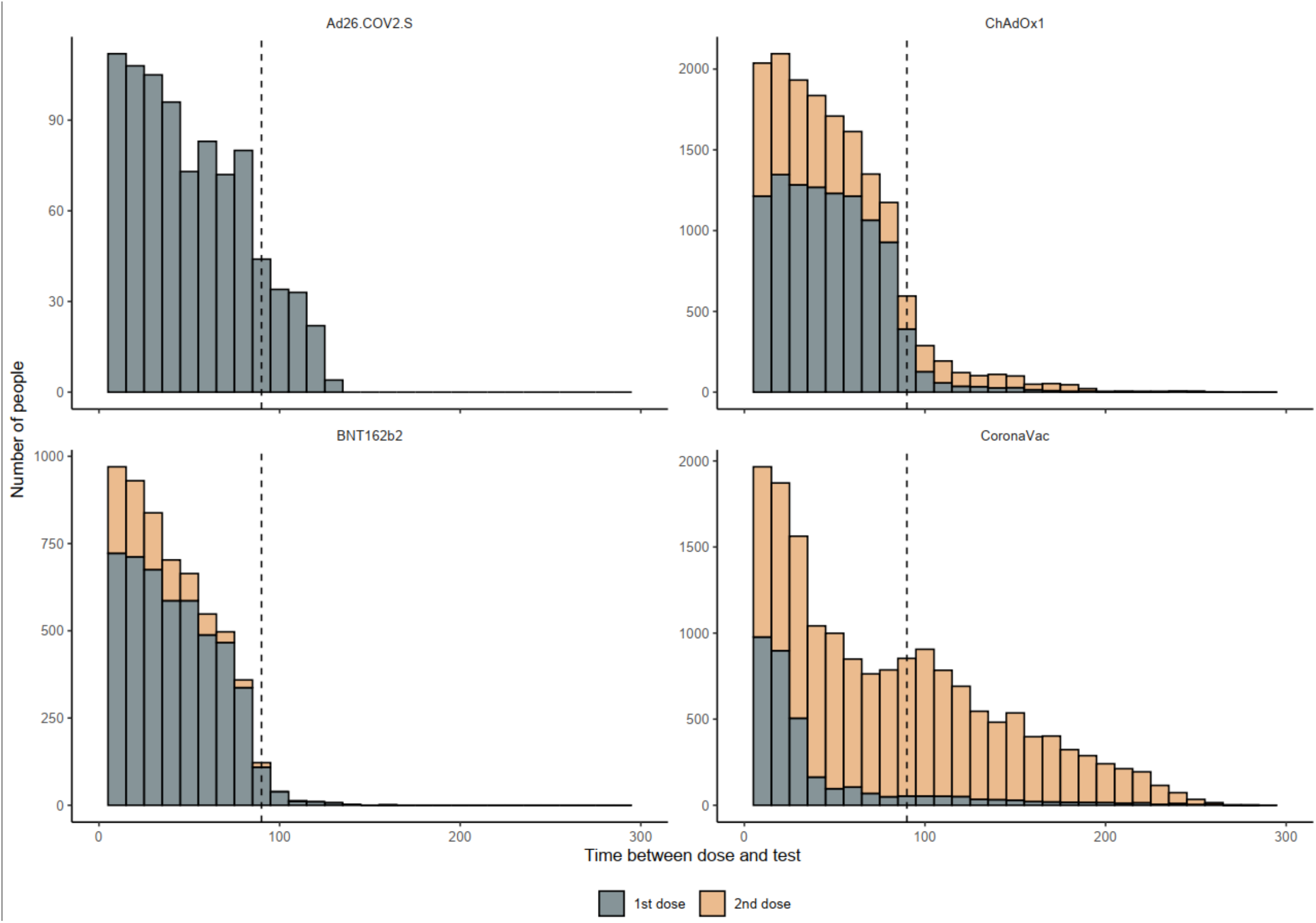
Time elapsed between the first (gray) or second (orange) dose and RT-PCR test among vaccinated people who had a prior SARS-CoV-2 infection. A dashed line is shown at 90 days.

**Figure A4.**
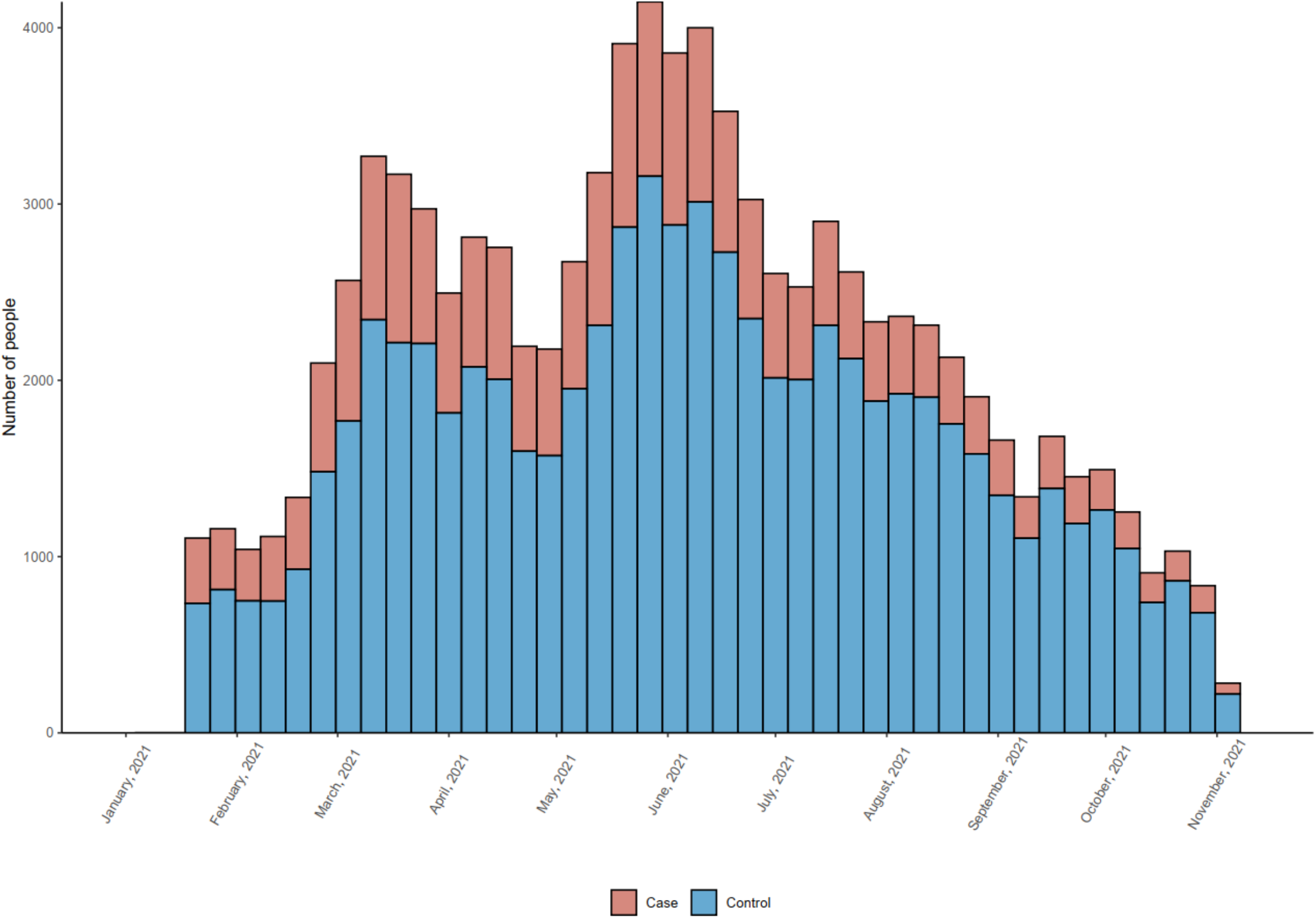
Number of enrolled discordant case-control pairs by date of RT-PCR testing during the study period between January 18 and November 11, 2021.

**Figure A5.**
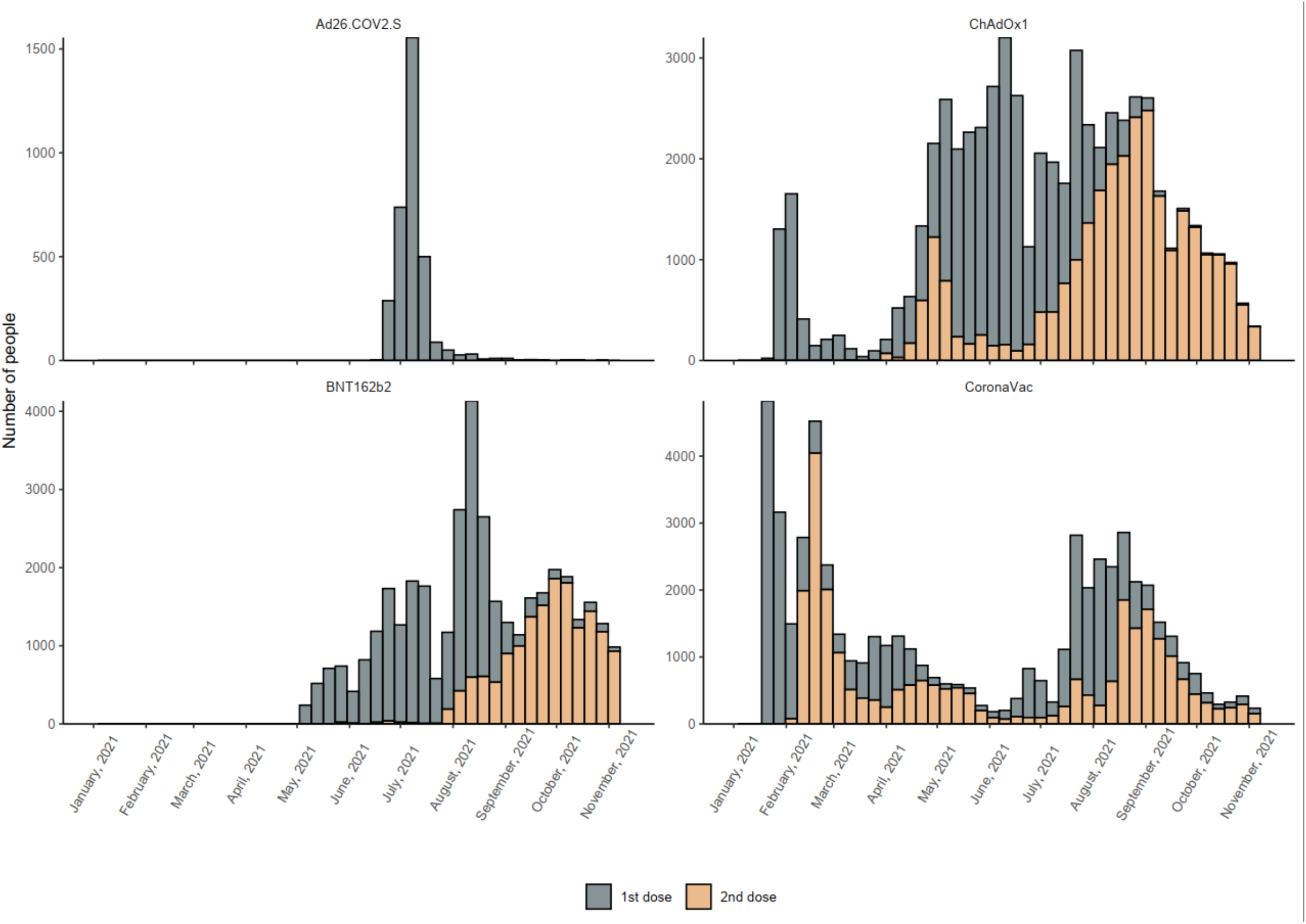
Week of vaccination for AD26.COV2.S, ChAdOx1, BNT162b2 and CoronaVac among matched participants.

**Table A1.** Characteristics and distribution of individuals according to vaccination status.

| **Characteristic** | **BNT162b2**<br>**N = 9,283** | **ChAdOx**<br>**N = 26,579** | **CoronaVac**<br>**N = 32,611** | **Ad26.COV2.S**<br>**N = 1,292** | **All vaccines**<br>**N = 69,765** | **Unvaccinated**<br>**N = 97,856** |
| --- | --- | --- | --- | --- | --- | --- |
| Case | 994 (10.7) | 3,114 (11.7) | 3,737 (11.5) | 155 (12.0) | 8,000 (11.5) | 14,566 (14.9) |
| Control | 8,289 (89.3) | 23,465 (88.3) | 28,874 (88.5) | 1,137 (88.0) | 61,765 (88.5) | 83,290 (85.1) |
| Individuals | 5,935 (63.9) | 15,897 (59.8) | 17,008 (52.2) | 877 (67.9) | 39,717 (56.9) | 51,275 (52.4) |
| Age (years) | 36 (28–44) | 40 (33–50) | 38 (31–45) | 40 (36–44) | 38 (31–47) | 35 (28–42) |
| Sex-female | 5,635 (60.7) | 17,547 (66.0) | 23,729 (72.8) | 734 (56.8) | 47,645 (68.3) | 56,917 (58.2) |
| Race |  |  |  |  |  |  |
| White | 4,120 (44.4) | 12,967 (48.8) | 15,325 (47.0) | 639 (49.5) | 33,051 (47.4) | 44,654 (45.6) |
| Mixed | 3,324 (35.8) | 8,650 (32.5) | 11,521 (35.3) | 402 (31.1) | 23,897 (34.3) | 34,889 (35.7) |
| Black | 511 (5.5) | 1,512 (5.7) | 1,770 (5.4) | 71 (5.5) | 3,864 (5.5) | 4,760 (4.9) |
| Indigenous or Asian | 362 (3.9) | 1,297 (4.9) | 1,976 (6.1) | 42 (3.3) | 3,677 (5.3) | 6,511 (6.7) |
| (Missing) | 966 (10.4) | 2,153 (8.1) | 2,019 (6.2) | 138 (10.7) | 5,276 (7.6) | 7,042 (7.2) |
| Region of residence |  |  |  |  |  |  |
| Central-west | 1,114 (12.0) | 3,222 (12.1) | 2,559 (7.8) | 186 (14.4) | 7,081 (10.1) | 8,218 (8.4) |
| North | 715 (7.7) | 1,399 (5.3) | 2,540 (7.8) | 47 (3.6) | 4,701 (6.7) | 9,541 (9.8) |
| Northeast | 1,408 (15.2) | 5,193 (19.5) | 8,219 (25.2) | 140 (10.8) | 14,960 (21.4) | 20,826 (21.3) |
| South | 1,104 (11.9) | 2,441 (9.2) | 2,680 (8.2) | 178 (13.8) | 6,403 (9.2) | 10,333 (10.6) |
| Southeast | 4,942 (53.2) | 14,324 (53.9) | 16,613 (50.9) | 741 (57.4) | 36,620 (52.5) | 48,938 (50.0) |
| Residence in state capital | 5,205 (56.1) | 13,194 (49.6) | 19,318 (59.2) | 576 (44.6) | 38,293 (54.9) | 47,887 (48.9) |
| Pregnancy | 145 (1.6) | 26 (0.1) | 110 (0.3) | 1 (0.1) | 282 (0.4) | 796 (0.8) |
| Post-partum | 7 (0.1) | 2 (0.0) | 12 (0.0) | 0 (0.0) | 21 (0.0) | 53 (0.1) |
| DM | 383 (4.1) | 2,007 (7.6) | 1,528 (4.7) | 28 (2.2) | 3,946 (5.7) | 3,582 (3.7) |
| Obesity | 245 (2.6) | 940 (3.5) | 845 (2.6) | 24 (1.9) | 2,054 (2.9) | 2,250 (2.3) |
| Immune supressed | 148 (1.6) | 456 (1.7) | 367 (1.1) | 17 (1.3) | 988 (1.4) | 1,348 (1.4) |
| CKD | 60 (0.6) | 261 (1.0) | 238 (0.7) | 1 (0.1) | 560 (0.8) | 536 (0.5) |
| Cardiac Disease | 702 (7.6) | 3,585 (13.5) | 3,042 (9.3) | 54 (4.2) | 7,383 (10.6) | 7,219 (7.4) |
| Medical comorbidities |  |  |  |  |  |  |
| 0 | 8,054 (86.8) | 21,031 (79.1) | 27,964 (85.8) | 1,195 (92.5) | 58,244 (83.5) | 85,991 (87.9) |
| 1 | 949 (10.2) | 4,096 (15.4) | 3,448 (10.6) | 74 (5.7) | 8,567 (12.3) | 9,252 (9.5) |
| ≥2 | 280 (3.0) | 1,452 (5.5) | 1,199 (3.7) | 23 (1.8) | 2,954 (4.2) | 2,613 (2.7) |
| Days, 1<sup>st</sup><br>positive test to<br>2<sup>nd</sup> test | 226 (161–323) | 238 (164–325) | 267 (191–338) | 225 (158–326) | 251 (175–332) | 203 (141–270) |
| Hospitalized<br>during 1st<br>infection | 432 (4.7) | 1,508 (5.7) | 1,612 (4.9) | 61 (4.7) | 3,613 (5.2) | 3,675 (3.8) |
| Days, 1st<br>vaccine dose to<br>2nd positive<br>test | 176 (112–276) | 166 (96–250) | 185 (99–242) | 186 (112–288) | 177 (100–248) | NA |
| Hospitalized<br>within 28 days | 183 (2.0) | 730 (2.7) | 712 (2.2) | 20 (1.5) | 1,645 (2.4) | 2,096 (2.1) |
| Death within<br>24 days | 16 (0.2) | 115 (0.4) | 165 (0.5) | 0 (0.0) | 296 (0.4) | 380 (0.4) |
| Hospitalized or<br>death | 184 (2.0) | 735 (2.8) | 729 (2.2) | 20 (1.5) | 1,668 (2.4) | 2,134 (2.2) |
| Vaccination<br>Status |  |  |  |  |  |  |
| One dose |  |  |  |  |  |  |
| 0-13 | 1,480 (15.9) | 2,551 (9.6) | 2,107 (6.5) | 193 (14.9) | 6,331 |  |
| ≥14 | 6,318 (68.1) | 15,813 (59.5) | 4,627 (14.2) | 1,099 (85.1) | 27,857 |  |
| Two doses |  |  |  |  |  |  |
| 0-13 | 448 (4.8) | 1,560 (5.9) | 2,362 (7.2) |  | 4,370 |  |
| ≥14 | 1,037 (11.2) | 6,655 (25.0) | 23,515 (72.1) |  | 31,207 |  |
| Days, 1st dose<br>to test | 35 (17–58) | 41 (21–64) | 20 (11–33) | 41 (20–70) | 34 (17–61) |  |
| Days, 2nd dose<br>to test | 23 (11–39) | 36 (18–66) | 76 (34–118) |  | 59 (27–105) |  |

Categorical variables reported as number (%). Numerical variables reported as median (IQR).

**Table A2.** Vaccine effectiveness among recently vaccinated individuals (0 to 6 days and 7 to 13 days post first dose).

|  | Symptomatic Disease |  | Hospitalization or Death |  |
| --- | --- | --- | --- | --- |
|  | Vaccine Effectiveness | 95% CI | Vaccine Effectiveness | 95% CI |
| **Unvaccinated** | Ref | - | Ref | - |
| **BNT162b2** |  |  |  |  |
| 0-6 days | -1.3% | -29.4 to 20.7 | -40.1% | -284.6 to 49 |
| 7-13 days | 27.2% | 9.8 to 41.2 | 29.4 | -73 to 71.2 |
| **ChAdOx1** |  |  |  |  |
| 0-6 days | 13.9% | -2.9 to 27.9 | -4.7% | -92.4 to 37.8 |
| 7-13 days | -25.5% | -45.5 to -8.2 | -48.4% | -131.1 to 4.8 |
| **CoronaVac** |  |  |  |  |
| 0-6 days | 21.0% | 2.3 to 36.1 | 8.7% | -77.0 to 52.9 |
| 7-13 days | -9.6% | -29.4 to 7.2 | 1.6% | -76.9 to 45.3 |
| **Ad26.COV2.S** |  |  |  |  |
| 0-6 days | 29.6% | -41.6 to 65.0 | -25.2% | -1817.7 to 91.8 |
| 7-13 days | -2.2% | -71.6 to 39.1 | -14.8% | -1045.6 to 88.5 |

**Table A3.** Vaccine effectiveness ≥14 days after series completion against symptomatic COVID-19 by subgroup.

| | 14-90 days | >90 days | p-value | 91-180 days | >180 days | p-value | 18-49 years | $\geq 50$ years | p-value |
| --- | --- | --- | --- | --- | --- | --- | --- | --- | --- |
| **BNT162b2** | 64.2%<br>(54.2-72.0) | 100%<br>(*) | 0.277 | 35.3%<br>(-2.7-59.3) | 70.7%<br>(61.1-77.9) | 0.011 | 64.6%<br>(53.0-73.3) | 66.9%<br>(46.0-79.7) | 0.891 |
| **ChAdOx1** | 55.5%<br>(50.5-60.1) | 56.8%<br>(46.6-65.1) | 0.544 | 47.0%<br>(32.7-58.3) | 58.4%<br>(53.7-62.6) | 0.201 | 54.8%<br>(49.4-59.7) | 59.9%<br>(50.8-67.3) | 0.274 |
| **CoronaVac** | 40.5%<br>(36.4-44.3) | 38.0%<br>(33.1-42.5) | 0.760 | 31.6%<br>(24.1-38.4) | 42.4%<br>(38.8-45.8) | 0.050 | 39.6%<br>(36.0-43.0) | 39.2%<br>(30.5-46.8) | 0.750 |
| **Ad26.COV2.S** | 46.1%<br>(32.7-56.7) | 30.6%<br>(-12.4-57.1) | 0.420 | 39.6%<br>(15.5-56.8) | 46.0%<br>(31.2-57.7) | 0.832 | 41.0%<br>(27.8-52.4) | 67.0%<br>(29.0-84.7) | 0.142 |

**Table A4.** Vaccine effectiveness ≥14 days after series completion against severe outcomes by subgroup.

|  | Vaccine waning<br>(time after series completion) |  |  | Natural immunity waning<br>(time after first infection) |  |  | Age |  |  |
| --- | --- | --- | --- | --- | --- | --- | --- | --- | --- |
| | 14-90 days | >90 days | p-value | 91-180 days | >180 days | p-value | 18-49 years | $\geq 50$ years | p-value |
| **BNT162b2** | 88.8%<br>(50.0-97.5) | 100%<br>(*) | 0.765 | 100%<br>(*) | 88.5%<br>(48.3-97.4) | 0.456 | 88.6%<br>(6.3-98.6) | 89.8%<br>(17.0-98.7) | 0.933 |
| **ChAdOx1** | 86.6%<br>(77.6-92.0) | 95.1%<br>(84.8-98.4) | 0.007 | 92.9%<br>(67.4-98.4) | 90.0%<br>(83.3-94.0) | 0.579 | 88.7%<br>(73.9-95.1) | 89.9%<br>(81.4-94.5) | 0.776 |
| **CoronaVac** | 86.6%<br>(79.8-90.3) | 74.4%<br>(63.3-82.2) | 0.012 | 74.1%<br>(57.9-84.1) | 84.10%<br>(78.4-88.4) | 0.118 | 84.9%<br>(76.6-90.3) | 79.2%<br>(69.7-85.7) | 0.153 |
| **Ad26.COV2.S** | 60.2%<br>(-10.8-85.7) | 41.0%<br>(-240.9-89.9) | 0.978 | 6.9%<br>(-179.6-69.0) | 81.8%<br>(17.1-96.0) | 0.064 | 65.2%<br>(3.3-87.5) | -32.3%<br>(-764.1-79.6) | 0.211 |

